# Synthesizing evidence from the earliest studies to support decision-making: to what extent could the evidence be reliable?

**DOI:** 10.1101/2022.03.20.22272675

**Authors:** Tianqi Yu, Lifeng Lin, Luis Furuya-Kanamori, Chang Xu

## Abstract

In evidence-based practice, new topics generally only have a few studies available for synthesis. As a result, the evidence of such meta-analyses raised large concerns. We investigated the robustness of the evidence of meta-analyses from these earliest studies. Real-world data from the Cochrane Database of Systematic Reviews (CDSR) were collected. We emulated meta-analyses with the earliest 1 to 10 studies through cumulative meta-analysis from eligible meta-analyses. The magnitude and the direction of meta-analyses with the earliest few studies were compared to the full meta-analyses. From the CDSR, we identified 20,227 meta-analyses of binary outcomes and 7,683 meta-analyses of continuous outcomes. Under the tolerable difference of 20% on the magnitude of the effects, the convergence proportion ranged from 24.24% (earliest 1 study) to 77.45% (earliest 10 studies) for meta-analyses of few earliest studies with binary outcomes. For meta-analyses of continuous outcomes, the convergence proportion ranged from 13.86% to 56.52%. In terms of the direction on the effects, even when only 3 studies were available at the earliest stage, the majority had the same direction to full meta-analyses; Only 19% for binary outcomes and 12% for continuous outcomes changed the direction as further evidence accumulated. Synthesizing evidence from the earliest studies is feasible to support urgent decision-making, and in most cases, the decisions would be reasonable. Considering the potential uncertainties, it is essential to evaluate the confidence of the evidence of these meta-analyses and update the evidence when necessary.

## Background

Systematic reviews and meta-analyses are a valid approach to provide comprehensive, transparent, and reliable evidence for better healthcare practice [1, 2]. In the hierarchy of evidence pyramid, high-quality systematic reviews and meta-analyses are at the top area over other types of evidence [3]. The high quality has been widely recognized in those systematic reviews with sufficient well-conducted studies for the meta-analyses. However, in practice, the median number of studies available for a meta-analysis is 3 (interquartile range [IQR]: 2 to 6) [4]. Our recent large-scale investigation of the Cochrane Database of Systematic Reviews (CDSR) also verified that more than 90% of the meta-analyses of healthcare interventions have less than 5 studies [5]. The small number of included studies in a meta-analysis may put the results at the risk of large fluctuations, which raises concerns about whether the results of such meta-analyses could be reliable to support the decision-making. This issue is even more critical for new topics (e.g., COVID-19), where only several earliest studies are available for the evidence synthesis.

To investigate this problem, several methodologists have examined the performance of meta-analyses based on the earliest few studies [6-11]. For example, in 2001, Ioannidis et al. [6] used a sample of 60 meta-analyses of randomized controlled trials in the field of pregnancy/perinatal medicine to assess the dispersion of relative changes of the effects of meta-analyses as evidence accumulated. They found that evidence based on the meta-analyses with 500 or fewer randomized patients should be interpreted cautiously, because the effects can easily be dissipated by future evidence. Trikalinos et al. [7] used a larger sample of meta-analyses (n=100) in mental health and obtained similar conclusions. Besides, Herbison et al. [9], Alahdab et al. [10], Wang et al. [11] conducted similar works; Table 1 presents a brief summary of similar important empirical investigations on this topic. These studies provide valuable information and guidance for evidence synthesis practice as well as decision-making.

**Table 1.**
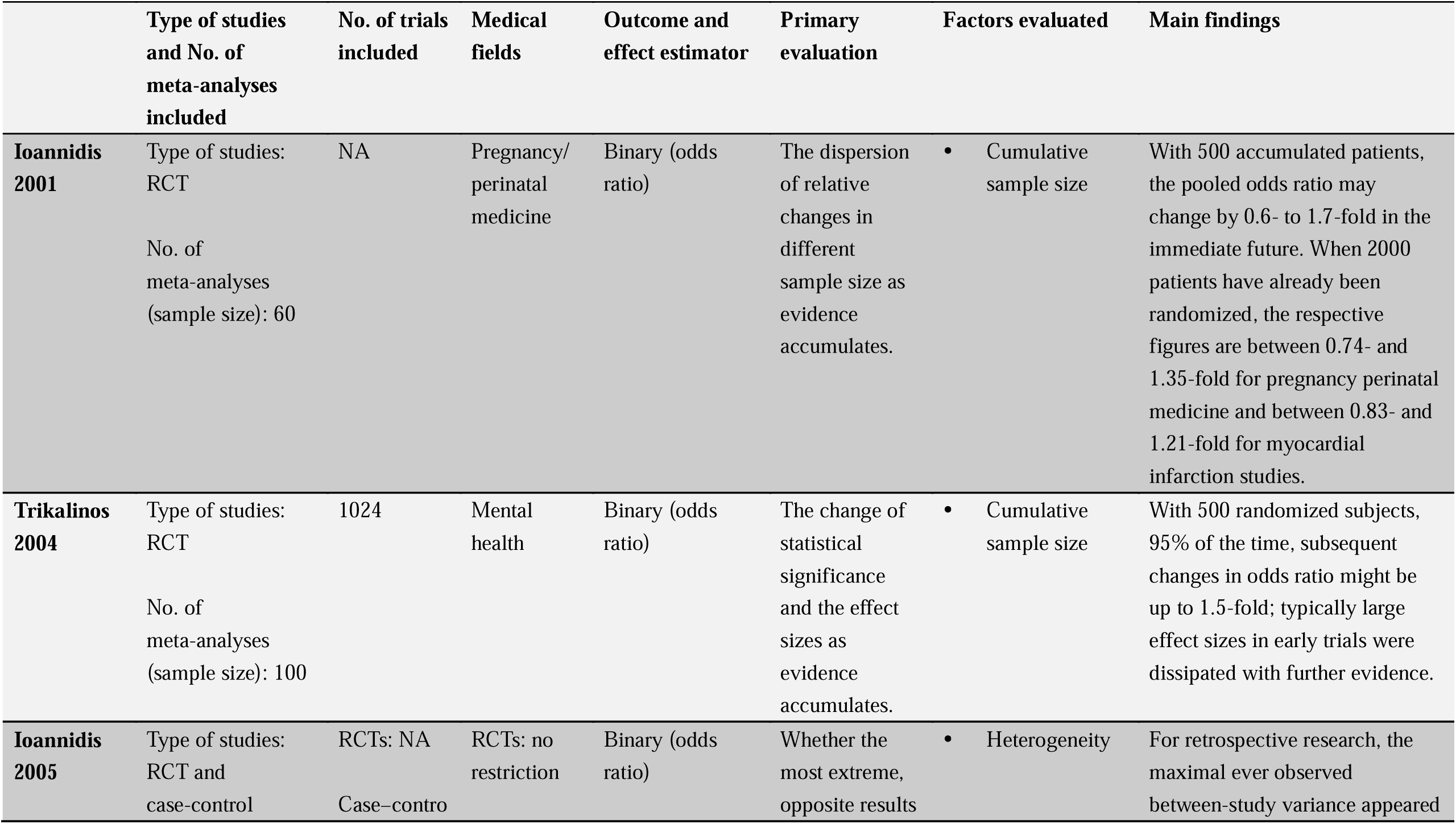
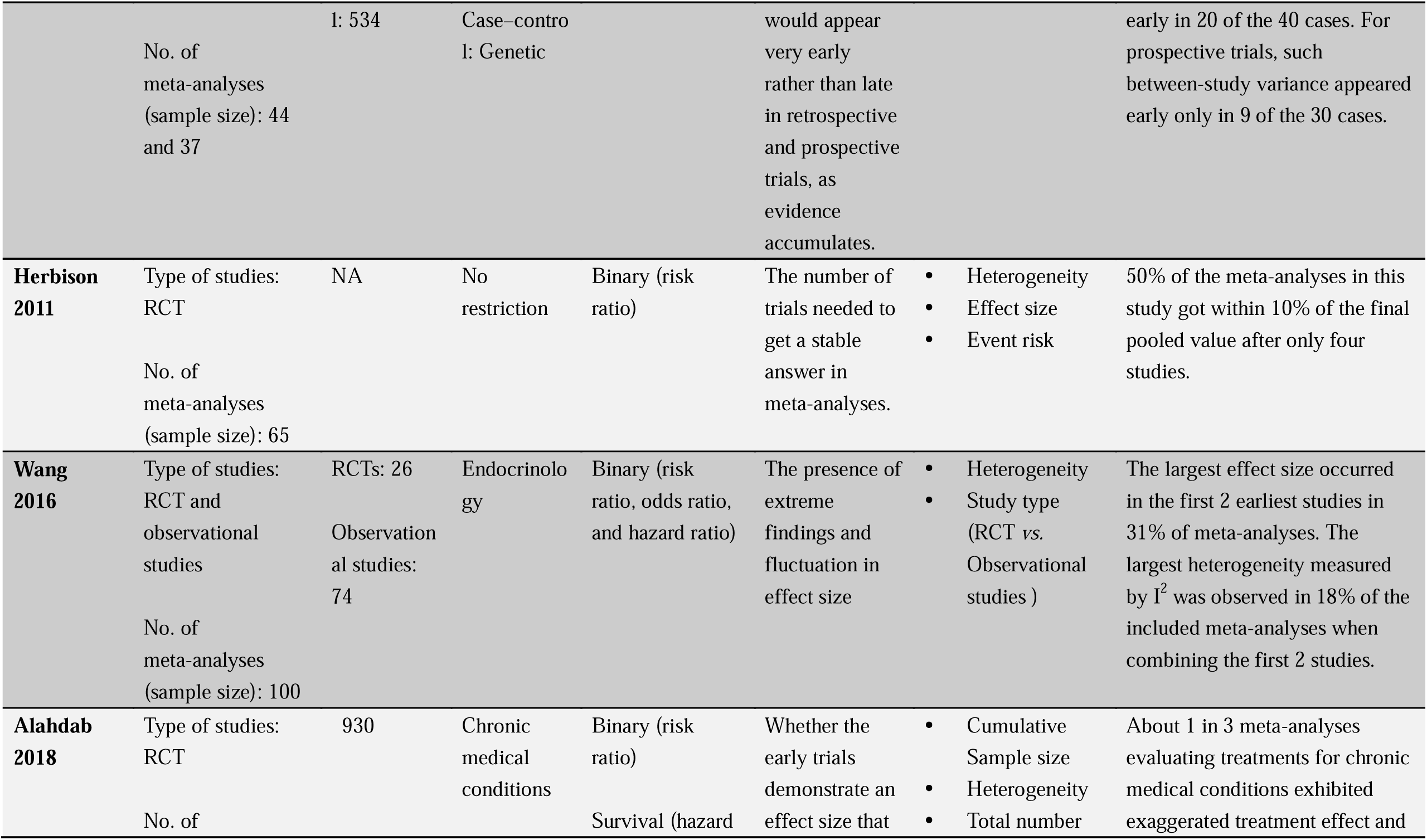
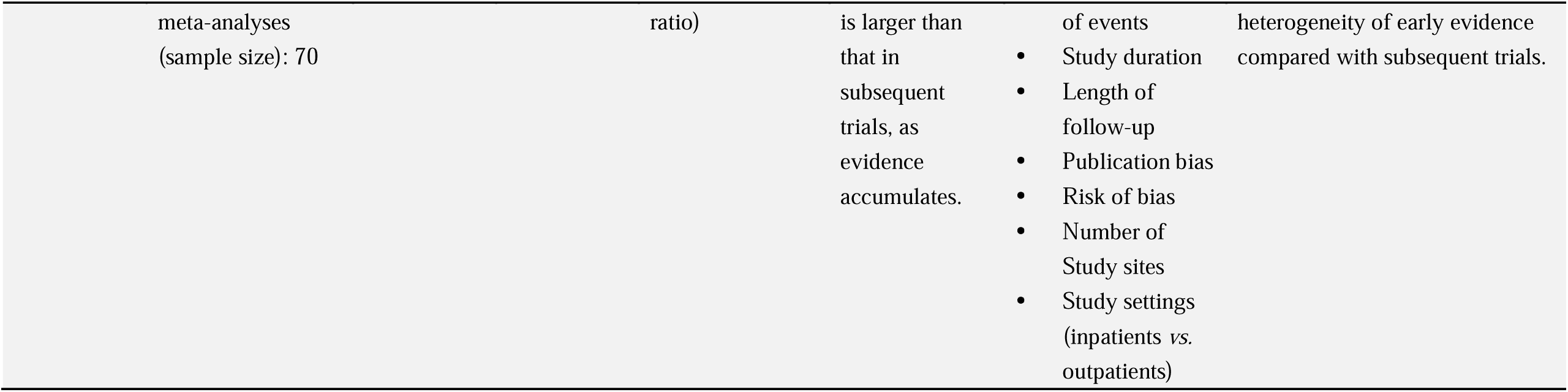
The basic information of the previous studies that investigated synthesizing evidence from the earliest studies.

Even so, these investigations are not absent of flaws. A common feature of these studies is that the sample size is small (ranging from 37 to 100); in addition, the topics considered in these studies were limited in one or two areas, which further limited the representativeness of the findings. Moreover, none of these studies investigated the robustness of the results for meta-analyses of continuous outcomes. Considering the potential limitations of previous studies, at least three questions need to be further addressed: 1) Under certain tolerances on the difference, for meta-analyses based on different numbers of earliest studies, how many of which the effects and *P*-values will change as evidence accumulates? 2) For different types of data under different effect estimators, do the extents of changes on the results from meta-analyses of earliest studies vary? 3) To what extent does the between-study variance, event risks, cumulative sample sizes, number of studies for final full meta-analysis, magnitude of effect size, as well as the publication bias affect the robustness of the results of meta-analyses based on earliest studies? These three questions eventually ended up with one question: *how to make reasonable decision-making based on the evidence of meta-analyses with earliest studies?*

In the current study, we utilized the large real-world dataset from the CDSR to evaluate the robustness of the evidence synthesized from early studies and attempted to address the above questions.

## Methods

### Data source

We collected real-world meta-analyses data from the CDSR. Considering that many Cochrane reviews published before 2003 were not available online and could only provide limited data, we utilized the data of Cochrane reviews published from January 2003 to May 2018 (accessed through Florida State University). This was done by using the R package “RCurl” [12] to automatically download the .rm5 files of these reviews automatically. Further, these .rm5 files were exported and saved as .csv files. Those Cochrane reviews without meta-analyses were identified and excluded at this stage. The lead authors then developed a Stata (Stata 14/SE, College Station, TX) program to clean the .csv files to make them suitable for analysis. The detailed process has been documented in our previous publications [5,13,14].

For the purpose of the current study, we only considered those meta-analyses with 5 or more studies [15]. These eligible meta-analyses were treated as full meta-analyses and were further used to emulate meta-analyses with the earliest few studies. This requires the information of publication year of each included study for the full meta-analyses, and therefore meta-analyses with missing data in this field were ineligible. In addition, we did not consider those meta-analyses with the total events count of zero in both arms because the effect of such meta-analyses would always be the same (i.e., risk difference = 0) that would overestimate the robustness.

The following information was of interest for the aim of the current study and was automatically collected by the Stata program: aggregate data of each study in each meta-analysis, publication year of each study, data types (e.g., binary, continuous), analytic models for meta-analyses (e.g., random-effects model), effect estimates, between-study variance estimates (i.e., I^2^ and tau^2^), and weight. For the binary outcomes, the effect estimates generally include the odds ratio (OR), risk ratio (RR), hazard ratio (HR), and risk difference (RD); for continuous outcomes, the effect estimates generally include the mean difference (MD) and standardized mean difference (SMD). Some information above was not relevant to the analysis but was important for the data cleaning process. For example, we used the weight and between-study variance estimates to help distinguish whether a dataset is a subgroup analysis or a new meta-analysis in the program.

### Emulating meta-analyses with the earliest few studies

In order to investigate the robustness of the results of meta-analyses based on the earliest few studies compared to the full meta-analyses, we employed a cumulative meta-analysis procedure with studies sorted by publication year within a full meta-analysis, from earliest to most recent. We considered the performance of the meta-analyses based on the earliest 1, 2, 3, 4, 5, 6, 7, 8, 9, and 10 studies. This decision was based on a recent survey that 87.6% of the meta-analyses within systematic reviews of COVID-19 had less than 10 studies [16].

We then compared the results of meta-analyses with the earliest studies to the meta-analyses with full sets of studies. It should be noted that some full-set meta-analyses contained no more than 10 studies, where it was impossible to consider all cases of including the earliest 1 to 10 studies. For example, when the full meta-analysis contained 7 studies, we could only consider including the earliest 1 to 6 studies. Therefore, for meta-analyses with 10 or fewer studies, we considered including the earliest 1 to *k*-1 studies, where *k* (≤10) represents the total number of studies in a full-set meta-analysis. Cumulative meta-analyses were performed using the “meta” package in R (See code in Appendix).

### Statistical analysis

For meta-analyses of binary outcomes, both the OR and RD were estimated. For meta-analyses of continuous outcomes, the SMD was estimated in all cases, while the MD was estimated only when the review authors used it in the original meta-analyses. Considering the potential between-study variance on the results, we employed two analytical models for all meta-analyses, i.e., the Knapp-Hartung (K-H) model [17] and inverse-variance heterogeneity (IVhet) model [18]. A recent simulation study has suggested that these two methods have the best performance for meta-analyses with few included studies [19]. A continuity correction of 0.5 was applied to all cells of studies with zero events in a single arm since the sample sizes of treatment and control arms were generally balanced [20]. For studies with zero events in both arms, there are currently some controversies of how to deal with such studies when using relative risk as the effect estimator [21, 22]. Since we used the standard inverse-variance weighted models where both effect and variance of such studies cannot be defined, we did not pool such studies in the meta-analyses (for OR) as suggested by Cochrane Handbook [23]. The I^2^ proposed by Higgins et al. [24] was used to measure the amount of heterogeneity for all cases.

The magnitude and direction of the effects were compared for meta-analyses of the earliest studies against the full meta-analyses. The significance of *P*-value was also compared with alpha = 0.05 as the level of significance. For the comparison of the magnitude, we used the absolute percentage *difference* of the effect of the meta-analysis with the earliest studies by treating the effect of the full meta-analysis as the “reference effect”. It was calculated by 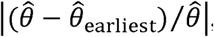, where 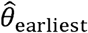 was the effect of a meta-analysis of the earliest studies and 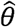 was the effect of the full meta-analysis. We set four cut-offs, 5%, 10%, 15%, and 20%, representing different tolerance levels for differences; when a difference was smaller than the certain cut-off, we treated it as convergent, and vice versa. The cut-off levels were based on a previous study, which documented that a median deviation of 10% (IQR: 5%-15%) was acceptable by methodologists [25].

For the comparison of the direction of the effects between meta-analyses with the earliest studies and full meta-analyses, we considered the following two situations. First, meta-analyses with the earliest studies showed the effect favouring the treatment group, while the full meta-analyses showed the effect favouring the control group, or vice versa. Second, meta-analyses with the earliest studies showed no effect (e.g., OR=1 or MD=0), while the full meta-analyses showed the effect favouring one of the groups, and vice versa. For meta-analyses with small effects, the direction was more likely to be changed, and mostly the change was of less interest because the bias was tolerable (e.g., OR_earliest_=1.01 vs. OR_full_=1.00). Therefore, as a sensitivity analysis, we further summarized the potential change of the directions of meta-analyses with the earliest studies vs. full meta-analyses for meta-analysis with a difference larger than 20% (the maximum tolerate cut-off of the current study).

We further evaluated the impact of event risks, effect sizes, heterogeneity, cumulative sample size, the total number of studies, and publication bias on the effects of meta-analyses with the earliest studies compared to full meta-analyses. We stratified meta-analyses with the earliest studies by each of the above factors and then summarized their convergence and direction-change proportion of the effects. First, the following groups were created in terms of the event risks of the control arm: 0-0.01, 0.01-0.05, 0.05-0.1, 0.1-0.2, 0.2-0.5, 0.5-0.75, and 0.75-1. For effect sizes, we divided meta-analyses with the earliest studies into three groups, i.e., “small effect” group, “moderate effect” group, and “large effect” group (see details in Appendix, Table S1). The definition was based on existing medical research guidelines [26] as well as the distribution of the effects by the current empirical dataset. For heterogeneity, we divided meta-analyses with the earliest studies into 5 groups according to their I^2^: I^2^=0 and I^2^ within 0-30%, 30%-60%, 60%-75%, and 75%-100%. For cumulative sample size, we classified meta-analyses with the earliest studies into the 4 groups according to previous studies [6, 7]: 0-499, 500-999, 1000-2000, >2000. For the total number of studies, the following groups were considered: 0-11, 12-20, 21-30, 31-50, >50. We set the first group as 0-11 instead of 0-10. This is because the full meta-analyses consist of 10 studies can only include the earliest 9 studies at most. If the first group was set to be 0-10, when the number of the included earliest studies was 10, the first group would be absent. The final stratification was done by whether there is publication bias, measured by the LFK index^27^ [27] (an LFK index between −1 and 1 means no publication bias, and vice versa).

## Results

From 2,693 Cochrane reviews, we identified 20,227 eligible meta-analyses of binary outcomes, including a total of 237,035 studies, and 7,683 eligible meta-analyses of continuous outcomes, including a total of 80,680 studies (Appendix, Figure S1).

### Meta-analyses with the earliest studies vs. full meta-analyses: magnitude

Figure 1 presents the comparisons of the magnitude between meta-analyses with the earliest studies against full meta-analyses. For binary outcomes, when measured with OR, the convergence proportion of meta-analysis with the earliest studies ranged from 6.64% (earliest 1 study) to 34.99% (earliest 10 studies) under the tolerance of 5%, 12.86% (earliest 1 study) to 54.57% (earliest 10 studies) under the tolerance of 10%, 18.94% (earliest 1 study) to 67.86% (earliest 10 studies) under the tolerance of 15%, and 22.24% (earliest 1 study) to 77.45% (earliest 10 studies) under the tolerance of 20%. The convergence proportion of meta-analyses with the earliest studies could reach 70% or above when the tolerance was set to 20% and 7 or more studies were available at the earliest stage. There was a very poor convergence (less than 40%) for the RDs, even if as many as 10 studies were available (Appendix, Figure S2-S5). For continuous outcomes, when measured by MD, the convergence proportion of meta-analyses with the earliest studies ranged from 3.59% (earliest 1 study) to 22.96% (earliest 10 studies) under the tolerance of 5%, 7.35% (earliest 1 study) to 37.12% (earliest 10 studies) under the tolerance of 10%, 10.88% (earliest 1 study) to 45.71% (earliest 10 studies) under the tolerance value 15%, and 13.86% (earliest 1 study) to 56.52% (earliest 10 studies) under the tolerance of 20%. The convergence proportion of meta-analyses with the earliest studies was almost below 50% in all cases for continuous outcomes (Appendix, Figures S2-S5). There was a slightly higher convergence proportion when measured by the SMD (Appendix, Figures S2-S5).

**Figure 1.**
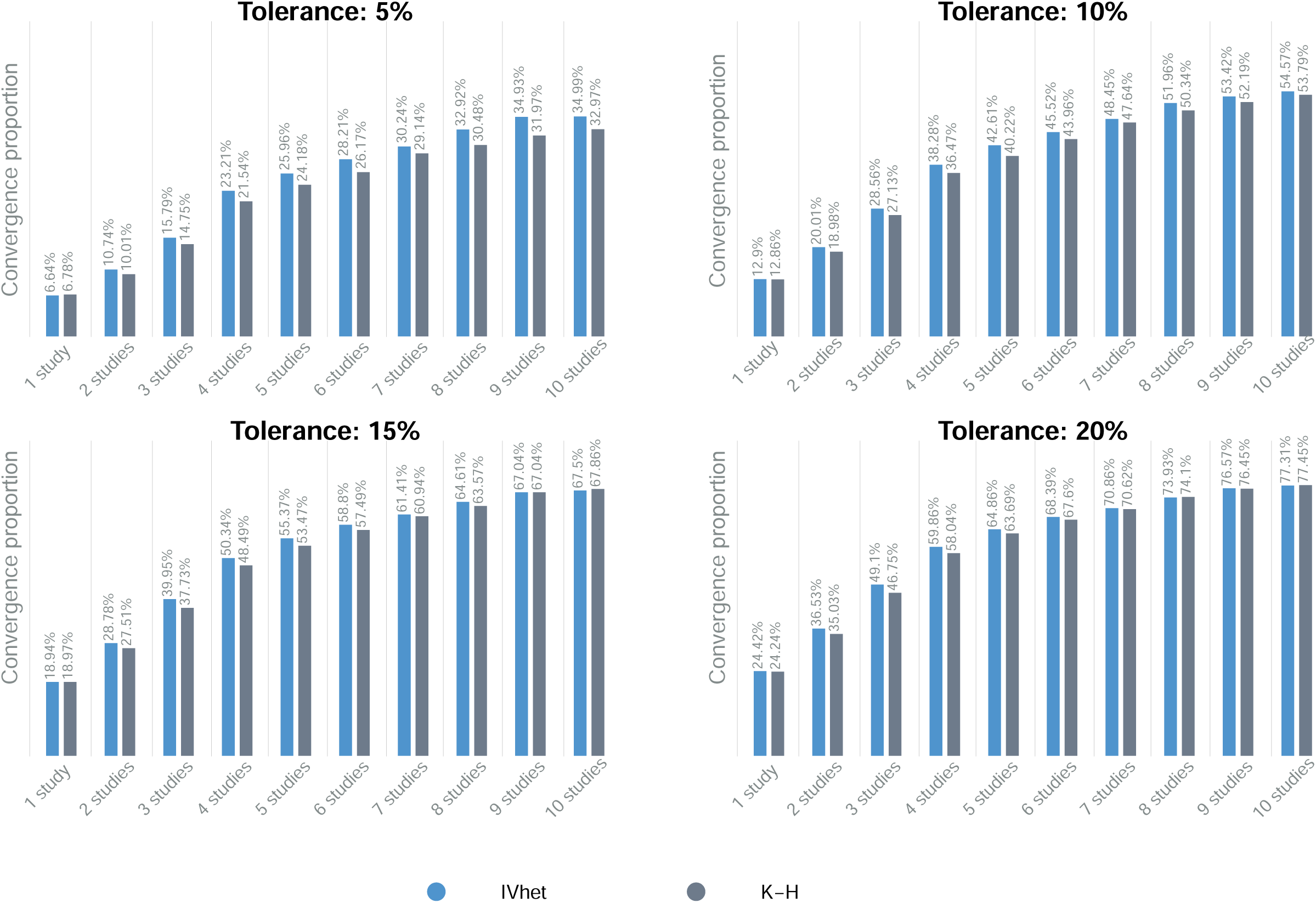
The convergence proportion under different tolerance values when the OR was used as effect estimator. *IVhet: inverse-variance heterogeneity model; K-H: Hartung-Knapp model*.

### Meta-analyses with the earliest studies vs. full meta-analyses: direction

Figure 2 presents the comparisons on the direction on the effects of meta-analyses with the earliest studies against full meta-analyses. For binary outcomes, the proportion of meta-analyses of the earliest studies with the effects (OR) in a different direction to full meta-analyses ranged from 8.84% (earliest 10 studies) to 27.59% (earliest 1 study). The RD showed a higher proportion. For continuous outcomes, the proportion of meta-analyses of the earliest studies with the effects (MD) in a different direction to full meta-analyses ranged from 2.23% (earliest 10 studies) to 23.02% (earliest 1 study), which were higher than SMD.

**Figure 2.**
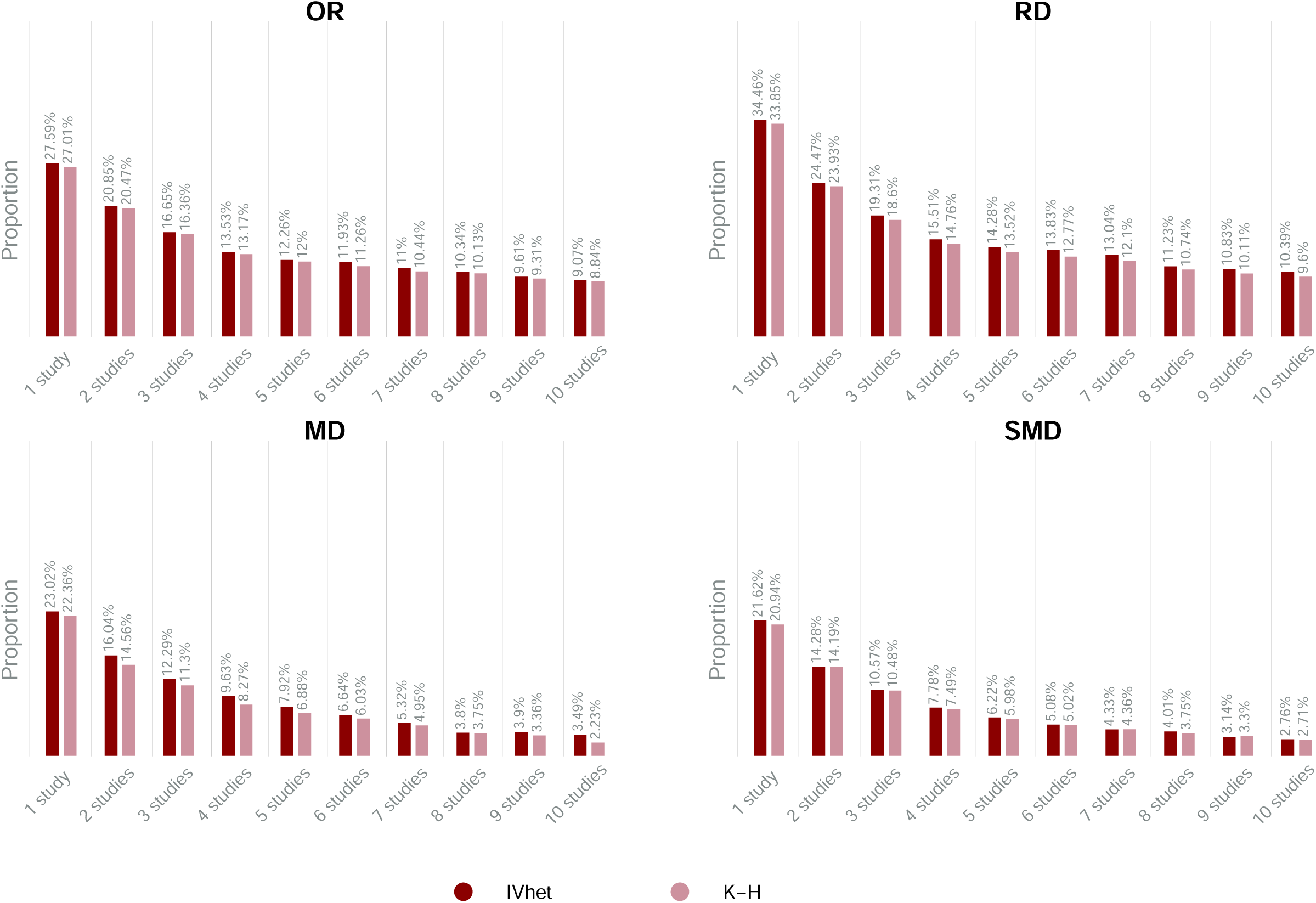
The proportion of meta-analyses of the earliest studies having a different direction of effect. *IVhet: inverse-variance heterogeneity model; K-H: Hartung-Knapp model*.

In our sensitivity analyses, for meta-analyses of the earliest studies with bias >20%, the proportion of the effects (OR) in a different direction to full meta-analyses ranged from 8.58% (including the earliest 10 studies) to 31.18% (including the earliest 1 study). Again, the RD showed a higher proportion of having a different direction. For continuous outcomes, the proportion of meta-analyses of the earliest studies with the effects (MD) in a different direction to full meta-analyses ranged from 2.60% (including the earliest 10 studies) to 26.35% (including the earliest 1 study).

### Meta-analyses with the earliest studies vs. full meta-analyses: significance

Figure 3 presents the comparison of the significance based on *P*-values of meta-analyses with the earliest studies against full meta-analyses. In general, compared to the effects, there was a higher proportion of *P*-value that changed the significance, which ranged from 12.69% to 47.68%. As more studies accumulated, fewer changed the significance. After including 5 or more studies, the proportion became steady at about from 14% to 20%. We put a special focus on the situation that meta-analyses with the earliest studies showed a significant effect (*P*<0.05) while the full meta-analyses showed non-significance (*P*>0.05). Our results suggested that when 2 or more studies were available at the earliest stage, only about 4% (K-H) or 7% (IVhet) of the meta-analyses with *P*-values changed from significant to non-significant when further studies were added (Appendix, Figure S6-A). For those with the earliest studies showed a non-significant effect (*P*>0.05), very few (0% to 0.16%) changed to a significant effect (*P*<0.05) when further studies were added (Appendix, Figure S6-B).

**Figure 3.**
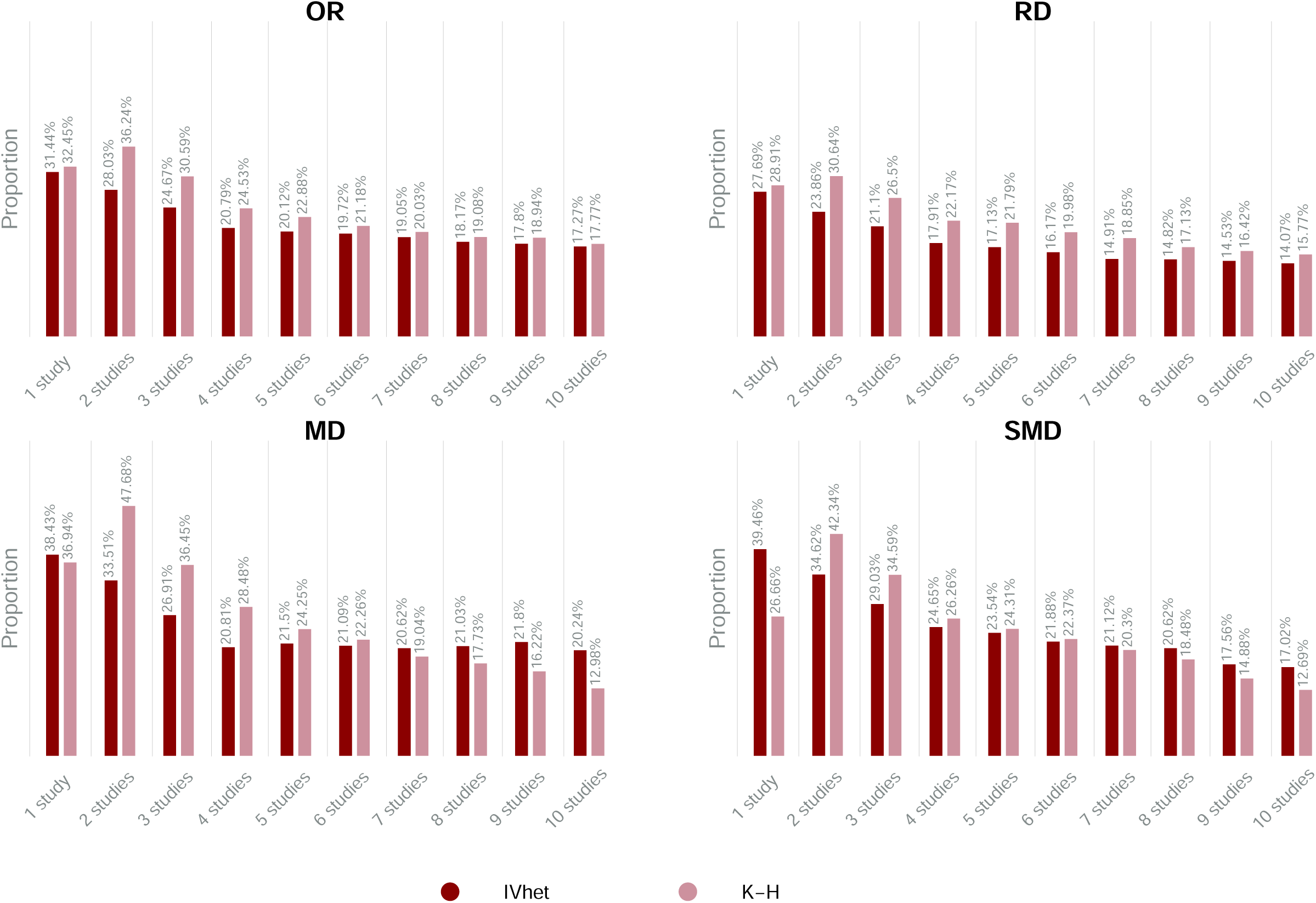
The proportion of meta-analyses of the earliest studies with significance changed in the full meta-analyses. *IVhet: inverse-variance heterogeneity model; K-H: Hartung-Knapp model*.

### Subgroup analysis

Subgroup analyses were conducted by different amounts of heterogeneity, publication bias, event risks, magnitude of the effects, cumulative sample size, and the total number of studies (Appendix, Table S2, Figures S7-S30). Our results suggested that for those meta-analyses of the earliest studies with high between-study heterogeneity, the convergence proportion was slightly higher than those with lower heterogeneity. Meta-analyses of the earliest studies with event risks less than 0.01 had a poor convergence in all cases, but the convergence became better as the event risk increased. However, for the magnitude of the effect, we observed that when measured by ORs, the convergence proportion of meta-analyses of the earliest studies with small effect sizes was more than three times higher than those with large effect sizes in some cases. Conversely, when measured by RDs, meta-analyses of the earliest studies in large effect sizes group had a higher convergence proportion than those in other groups. For meta-analyses with continuous outcomes, the convergence proportion of meta-analyses with the earliest studies was the highest when effect sizes were moderate. Meta-analyses of earliest studies with large sample sizes had higher convergence proportions than those with small sample sizes, and sample sizes had a more substantial impact on convergence proportions when measured by ORs than by other effect estimators. Only when the total number of studies of full meta-analyses was below 10, meta-analyses of the earliest studies had a considerably higher convergence proportion, except for those measured by MDs. Publication bias measured by the LFK index seemed to have no obvious impact on convergence.

In terms of direction, meta-analyses based on the earliest studies with zero between-study heterogeneity, lowest event risks, smallest effect sizes, and cumulative sample sizes less than 500 had a considerably higher proportion of effects in a different direction to full meta-analyses than other groups (Appendix, Figures S31-S36). For other groups, as between-study heterogeneity, event risks, effect sizes, and sample sizes increased, the proportion of meta-analyses of earliest studies with effects in a different direction decreased slightly. For binary outcomes, meta-analyses of the earliest studies had the highest different direction proportion when the total number of studies in full meta-analyses was above 50. However, this was not the case for continuous outcomes. Again, publication bias had no obvious impact on direction.

When it comes to *P*-value, before including 4 or 5 studies, meta-analyses of the earliest studies with larger effect sizes, higher event risks, and smaller sample sizes had a higher proportion of *P*-values changed the significance to the final pooled *P*-values (Appendix, Figures S37-S42). After including 5 or more studies, no obvious regularity of change in such proportion showed up. Contrary to the impact of heterogeneity on the effect direction, meta-analyses of the earliest studies with the largest I^2^ had the highest proportion of *P*-values changed the significance to full meta-analyses. Compared to continuous outcomes, the total number of studies in full meta-analyses had a greater impact on binary outcomes. The proportion of meta-analyses of the earliest studies with change in significance decreased as the total number of studies increased.

### Example from the CDSR

We used a meta-analysis [28] from the CDSR as an example to show the robustness of the effects by using the earliest studies. The meta-analysis investigated the efficacy of endothelin receptor antagonists (ERAs) in pulmonary arterial hypertension (Appendix, Table S3), with 17 trials involving a total of 3,322 participants. The improved functional class as a primary binary outcome in the meta-analysis was selected as an example. Among the 17 trials, two that did not report the selected outcome were excluded. For the remaining 15 trials, we performed meta-analyses with the earliest 1 to 14 trials separately, and compared the results of these meta-analyses to the full meta-analysis under the K-H model. Our results suggested that meta-analyses with the earliest 1 to 14 studies showed the same direction of ORs to the full meta-analysis (favoring the ERAs treatment). After including the earliest 7 or more studies can mostly reach around the final pooled value (Appendix, Figure S43). In this example, even if we used the earliest studies, we could mostly make a reasonable decision, at least in terms of the effect direction.

## Discussion

In this study, we investigated the robustness of the evidence from the earliest studies for meta-analysis by using a large real-world dataset. Our findings suggested that, under a 20% tolerance of the difference and when there were 7 or more studies available at the earliest stage, the effects of 70% meta-analyses of binary outcomes could be robust in terms of the magnitude. For meta-analyses of continuous outcomes, only up to 50% with the effects could be robust in terms of the magnitude. However, when considering the direction of the effects, as long as 3 studies were available at the earliest stage, the direction of 81% meta-analyses of binary outcomes and 88% meta-analyses of continuous outcomes would not be changed. In addition, when 4 or more studies were available at the earliest stage, the significance of *P-*value in at least 80% of the meta-analyses would not change in the future.

We also found that for binary outcomes, using the OR as the effect estimator is better than the RD to establish the earliest evidence from a meta-analysis in terms of robustness. This could be partly explained by the nature of these two measurements that the OR is “portable” while the RD is not [29]. Specifically, when new evidence is added, the RD would be largely impacted by the different baseline risks and thus becomes unstable, but this is not the case for the OR. In addition, previous studies have recorded that the RD tends to be more heterogeneous and has lower statistical powers than the OR in meta-analyses [30, 31]. These limitations of the RD support that the OR is a better effect estimator than the RD for meta-analyses with the earliest studies.

Our subgroup analysis suggested that, for meta-analyses with larger amounts of heterogeneity, the effects were less likely to be changed in terms of the magnitude and direction than those with smaller heterogeneity. This seems not intuitive, but one reason may be that meta-analyses with small amounts of heterogeneity are more likely to be meta-analyses with rare events (Appendix, Table S4)—subgroup analysis by event risk of the current study suggested that meta-analyses of rare events are much less robust than those with common events. Another reason could be that meta-analyses with small I^2^ included a higher proportion of small individual studies (Appendix, Table S5), adding to the instability of meta-analyses [32]. We also compared the I^2^ in each iteration in the cumulative meta-analysis procedure, and it generally remained stable and irrelevant to the additional accumulated studies (Appendix, Table S6). This observation warrants further in-depth investigation of the mechanisms.

As shown in our sensitivity analyses, by different extents of tolerance on the difference, the robustness of the results of meta-analyses with the earliest studies depended largely on the tolerance. Low tolerance on the difference (e.g., 5%) of meta-analyses with the earliest studies means less convergence, and vice versa. When decision-makers need accurate estimation (with a small difference) to support their decision, they will face the risk that the evidence of the earliest studies could be likely unstable. In practice, low tolerance is unrealistic for urgent decisions; the most important and urgent task is to predict the direction of the effects. Therefore, when the prediction on the direction of the effect is correct, we can make a reasonable decision. In the current study, we found that even only 3 studies were available at the earliest stage, the majority of the meta-analyses would properly predict the direction of the effects. This finding suggested that using the earliest evidence to support decision-making is valid and reasonable in most situations. It also may have some implications for decision-making in the pandemic of COVID-19 or other urgent public health events. On the other hand, the findings of the current study may indicate potential research waste in related topics—when there is robust and consistent evidence, perhaps further studies are no longer needed. However, it remains challenging to decide when the evidence is robust and conclusive.

### Implications for decision-making

As presented, there were up to 19% of the situations for meta-analyses of binary outcomes and 12% meta-analyses of continuous outcomes where conclusions changed when new evidence was included. Such uncertainty and its susceptibility to other factors further highlight the need for deliberation in decision-making. In addition, current researchers largely rely on the significance of *P*-value or whether the confidence interval contains the null effect for the inference, which is prone to misleading decisions [33]. There are some practical suggestions for systematic review authors to form a reasonable conclusion for decision-makers.

- First, it is not recommended to rely only on the significance of the *P*-value to form the conclusions. As pointed out by the American Statistician Association, “*P-values do not measure the probability that the studied hypothesis is true, or the probability that the data were produced by random chance alone*.” [34] In addition, *P*-value is sensitive to sample size, which has been criticized by previous studies [35]. Unlike *P*-value, the effect size is less likely to be influenced by sample size. Our results also support that the direction of the effects was less likely to change than the significance of *P*-values as new evidence accumulated. The confidence interval also provides a way to measure compatibility [36]. Nevertheless, it should be noted that some tests against chance are still important, as pointed out by John et al [37].
- Second, it is not recommended to use the RD as an effect estimator for meta-analyses of binary outcomes when only a few earliest studies are available; instead, the OR is a better option. Similarly, the SMD could be a better choice for continuous outcomes in such a situation.
- Third, assumption-free methods, such as the IVhet and K-H models, should be considered as the primary choice [17]. These methods do not rely on the normal distribution assumption for heterogeneous studies and could provide better parameter estimation, especially when only a few studies are available. At the same time, researchers should avoid using the conventional random-effect model, i.e., DerSimonian-Laird model [38], because well-established evidence suggested that this method showed poor performance for meta-analyses with a small number of studies [19].
- Fourth, meta-analysts may consider reporting prediction intervals [39] as they provide the range of the effect of a future new study to aid decision-making.
- Fifth, updating the evidence regularly when current evidence is insufficient to support reasonable decision-making.
- Last but not least, it is essential to evaluate the certainty of the evidence for each meta-analysis. Several tools are available, such as the Grading of Recommendations Assessment, Development and Evaluation (GRADE) [40]. This step also provides the basis for the judgment of whether a further update of the evidence is needed.

To the best of our knowledge, this is currently the largest empirical investigation for the robustness of the evidence produced by the earliest studies. The findings of the current study are expected to be useful to aid decision-makers in better forming the decision based on limited evidence and help future review authors better conduct meta-analyses at the initial stages of evidence evolution. In addition, current findings provide useful information for methodologists to develop a framework to rank the confidence of the earliest evidence.

Some limitations should be highlighted. First, about 8% of the samples were excluded from our analyses because these meta-analyses did not contain information of publication years of included studies. These can be considered as missing data, but they would have little impact on the results since the missing mechanism was likely at random [41] and the proportion of missing data was small. Second, the current study treated the results of full meta-analyses as the reference effect, and the analyses on the robustness were all based on this assumption. This is reasonable since it is the typical case in practice. However, even for the full meta-analyses, some of them are inevitably unstable due to the potentially low statistical power, limited sample size, low incidence of the outcomes, high risk of bias of included studies, or high between-study heterogeneity. Therefore, the uncertainty of the full meta-analyses would impact the findings of the current study. To reduce the impact of such uncertainty on our study, we removed those meta-analyses with less than 5 studies and employed three meta-analytic methods for the analyses. Even so, the findings should be interpreted with caution. Third, the current study was based on the CDSR, in which the systematic reviews were expected to have rigorous designs and implementations. However, the literature contains much more non-Cochrane reviews, among which many face serious methodological weaknesses; such weaknesses would impact the confidence of the results. Therefore, it is also recommended to assess the methodology rigorous of these meta-analyses with the earliest studies for better decision-making. Last, even if there were no systematic differences at all between earlier and later studies, the difference would still be positive in expectation because it is based on the absolute value of a difference between two quantities subject to random errors, and a difference between these quantities is virtually inevitable. Furthermore, this difference will be larger on average when the number of early studies included is smaller, because there will be more random errors in such cases.

## Conclusions

In conclusion, based on the findings of our empirical investigation, the utilization of the earliest studies in systematic reviews and meta-analyses to support decision-making is reliable in most situations. In more than 81% of the situations of binary outcomes, and 88% of the situations of continuous outcomes, the decisions would be reasonable (with the correct effect direction) based on the evidence from the earliest 3 or more studies. As more studies become available, the evidence will be more robust. However, the evidence synthesized from only the earliest 1 or 2 studies is likely to encounter large fluctuations in the future, as new studies appear. The event risks, magnitude of the effects, between-study heterogeneity, and sample sizes have some impact on the robustness of the results that should also be accounted for in the decision-making process. However, there are still considerable uncertainties, and thus it is essential to evaluate the confidence of the evidence of these meta-analyses for decision-making; in addition, it is also important to update the evidence when necessary.

### Highlights

1. ***What is already known***
  - New topics generally only have a few studies available for synthesis. As a result, the evidence of such meta-analyses raised large concerns.
2. ***What is new***
  - Under a 20% tolerance of the difference and when there were 7 or more studies available at the earliest stage, the effects of 70% meta-analyses of binary outcomes could be robust in terms of the magnitude. The direction of 81% meta-analyses of binary outcomes and 88% meta-analyses of continuous outcomes would not be changed based on the evidence synthesized from the earliest 3 or more studies.
3. ***Potential impact for Research Synthesis Methods readers outside the authors’ field***
  - The utilization of the earliest studies in systematic reviews and meta-analyses to support decision-making is reliable (with the correct effect direction) in most situations. The event risks, magnitude of the effects, between-study heterogeneity, and sample sizes have some impact on the robustness of the results. Considering the potential uncertainties, it is essential to evaluate the confidence of the evidence of these meta-analyses and update the evidence when necessary.

## Data Availability

All data produced in the present study are available upon reasonable request to the authors

## Declarations

### Consent for publication

Not applicable

### Ethics approval and consent to participate

Not applicable

## Acknowledgments

We would like to thank Prof. Suhail Doi from Qatar University for carefully reviewing the draft of this manuscript.

## Availability of data and material

We have no copyright to share the data with the public. For researchers wishing to obtain data for academic use, they are advised to contact the corresponding author.

## Authors’ contributions

CX conceived and designed the study; CX and LL collected the data; TY and CX analyzed the data and drafted the manuscript; LL and LFK provided methodology guidance, comments, and edits for the manuscript. All authors approved the final version for publication.

